# Efficacy of therapies in the treatment of Guillain-Barre Syndrome: a network meta-analysis

**DOI:** 10.1101/2020.06.03.20121780

**Authors:** Jingfeng Lin, Qiang Gao, Kang Xiao, Danfeng Tian, Wenyue Hu, Zhenyun Han

## Abstract

**Background:** Guillain-Barre syndrome (GBS) is an acute, paralyzing, inflammatory peripheral nerve disease. For treatments of the Guillain-Barre Syndrome, there are many kinds of therapies for this diseases. For comparing all of the therapies, such as immunoglobulin, plasma exchanging, etc. in the treatment of Guillain-Barre Syndrome(GBS) to better inform clinical practice, we used Network meta analysis to get the outcome of the Guillain-Barre Syndrome. The protocol had been submitted to PROSPERO:CRD: 42019119178.

**Methods:** Web of Science, PubMed, Embase, and the Cochrane library were searched for related articles. We identified citations and of these included 28 trials comprising 2474 patients and control group human beings. Network meta-analysis (NMA) was performed with two kinds of outcomes. We carried on R software with gemtc package and JAGS software to calculate results for different therapies. The consistency of direct and indirect evidence was also assessed by R software.

**Results:** Concerning for two outcomes, there were no improvement observed in methylprednisolone and prednisolone compared with placebo. PE and IVIg were illustrated to be effective over Placebo. There was no significant difference between different doses and times of PE and IVIg. On consistency examination between direct and indirect evidences, there were no obvious heterogeneity between all of therapies. Funnel plots indicates the possibility of publication bias in this study is small.

**Conclusion:** PE or IVIg have a significant efficiency for GBS patients. The effect of some combination treatments should be further explored. Corticosteroids have no significant effects on GBS.

## 1. Introduction

Guillain-Barre Syndrome (GBS) is a demyelinating polyradiculoneuropathy with an acute paralyzing disorder, typically symmetric, ascending and areflexia[1, 2]. Incidence varies between 0.66 and 1.79 cases per 100 000 persons in general population[2, 3]. It is an autoimmune disease caused by an immune reaction against an infectious agent that shares an antigen with nerves[4], This pathological mechanism has led to the use of immune therapies for Guillain-Barre syndrome. The common treatments for Guillain-Barre syndrome includes plasma exchange[5], intravenous immunoglobulin[6] and Corticosteroids[7]. There are many other ways as combination of therapies, non-routine dose and courses of IVIg(intravenous immunoglobulin), and unconventional treatments[8]. Many clinical trials have been conducted to investigate their therapeutic effects on Guillain-Barre Syndrome[9–12]. However, sample sizes of previous studies were relatively limited, and many kinds of therapeutic effects have not been compared with each other, as well as the assessment of Guillain-Barre Syndrome were not in consistence with each other. Thus, a network meta-analysis was needed to help carry out comparative analysis on the efficiency of therapies on Guillian-Barre Syndrome from previous trials with 2 endpoints. Network meta-analysis (NMA) is a statistical technique that allows comparison of multiple treatments in the same meta-analysis simultaneously. NMA can be performed under a frequentist or a Bayesian framework.[13] We carried on R software with gemtc package and JAGS software to calculate the rank probabilities of different therapies on two outcomes. The protocol has been submitted to PROSPERO:CRD: 42019119178.

## 2. Material and methods

### 2.1 Search strategy

We searched for Web of Science, PubMed, Embase, and the Cochrane library for related articles concerning the therapeutic effects of therapies for Guillian-Barre Syndrome. All therapies were enrolled, including PE, IVIg with different dose and courses, Corticosteroids, CSF filtration, combination of therapies, etc.. Articles published between January 1, 1980 and January 1, 2019 were retrieved in the search. The following Mesh terms and their synonyms and abbreviations were used to find relevant studies: “Guillain-Barre syndrome”, “polyradiculoneuropathy”, “polyneuropathies”, “methylprednisolone”, “prednisolone”, “IVIg or intravenous immunoglubin”, “plasma exchange”, etc.. Two authors independently screened titles and abstracts of retrieved articles to evaluate their qualification according to the inclusion criteria. Reference list of enrolled articles were also reviewed manually to improve the integrity of this study. This analysis was performed in accordance with Preferred Reporting Items for Systematic Reviews and Meta-Analyses (PRISMA) guideline. [14]

### 2.2 Evaluated outcomes and inclusion criteria

In study, Disability scale grade change after 4 weeks(outcome①) and rates of improvement by ≥1 grades of Disability scale after 4 weeks(outcome②)were applied to evaluate the on patients. The Disability scale has some version, all versions is approximately divided to seven layers: 0. Healthy. 1. Minor symptoms or signs of neuropathy but capable of manual work. 2. Able to walk without support of a stick but incapable of manual work. 3. Able to walk with a stick, appliance, or support. 4. Confined to bed or chair bound. 5. Requiring assisted ventilation.6. Dead. We included all randomized controlled trials (RCTs) or cohort trials, and included children and adults with GBS of all degrees of severity. We defined GBS according to internationally accepted diagnostic criteria[4] as acute polyradiculoneuropathy causing progressive weakness of two or more limbs, an onset phase not more than four weeks, reduced or absent tendon reflexes, and lacking alternative causes. We included studies that did not conform exactly to these criteria provided that the authors regarded GBS or one of its synonyms, such as acute idiopathic neuropathy or acute inflammatory demyelinating polyradiculoneuropathy, as the preferred diagnosis.

### 2.3 Data extraction

Two authors extracted relevant data from eligible articles independently. In the current study, information as follows were extracted: last name of first author, year of publication, origin country, type of clinical trial, number of subjects, treatments, outcomes. A 3rd author would resolve discrepancies after discussion. Disability grade change after 4 weeks was considered as the primary outcome in this study.

### 2.4 Statistical analysis

A network meta-analysis was performed in order to evaluate the efficacy of different treatments on GBS. Consequently, Bayesian network meta-analysis (NMA) was performed with a consistency model using getmc[15] package and JAGS software in R software to compare direct and indirect evidence. Moreover, the convergence graph, forest graph, density graph and heterogeneity test between direct and indirect evidence would be painted or assessed by R software. Besides getmc package, BUGSnet[16], netmeta [17], and dmetar[18] packages were also be required for making League Table Heatmap, Funnel plot, Direct evidence plot and Heterogeneity analysis heatmap. We set the parameter of bayes iterations as n.adapt = 5000, n.iter = 20000 to ensure the convergence.

## 3. Results

### 3.1 Study characteristics

A total of 2474 subjects from 28 trials were involved to investigate the efficacy of 15 kinds of therapies for Guillain-Barre Syndrome. Following Figure in Fig. 1 illustrated the process of study selection. Therapies involved in this meta-analysis were Plasmaexchange(PE), Intravenous immunoglobulin(IVIg), methylprednisolone(MTP), prednisolone(Pred), immunoabsorption plasmapheresis(IAPP), IFN-b 1a, BDNF(brain-derived neurotrophic factor), CSF filtration(CSF filter), Tripterygium polyglycoside(TWP), PE followed by IVIg(PE+IVIg), immunoabsorption followed by IVIg(IAPP+IVIg), IVIg (LFB) 0.4 g/kg/day for 3 days(Half dose of IVIg), IVIg 1 g/kg daily(twice dose/day of IVIg), Half-course of treatment of PE and IVIg +eculizumab.

**Figure 1.**
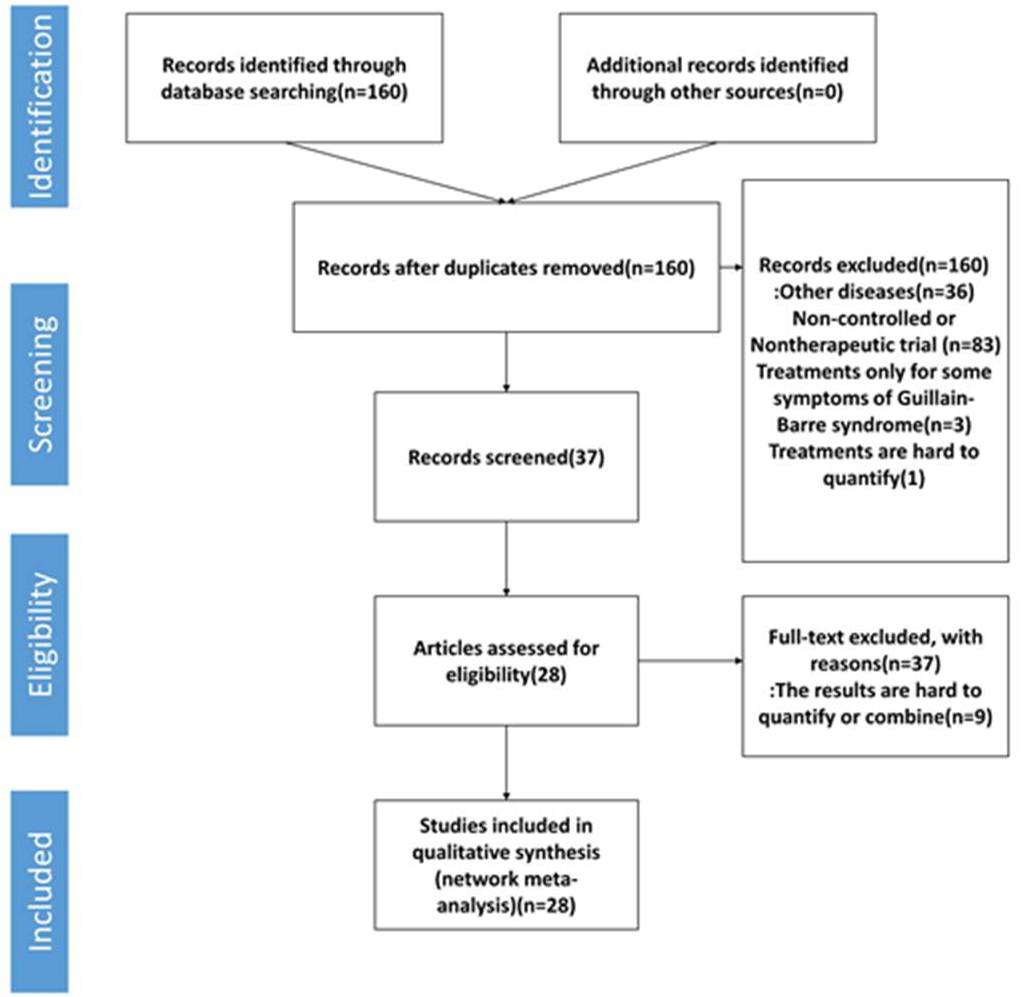
Flow chart of the search for eligible studies

### 3.2 Quality assessment

27 of 28 trials mentioned randomization, 19 RCTs (68%) used a specific random sequence generation method. Most of these RCTs(16 RCTs) mentioned allocation concealment. Only 10 RCT (36%) referred to the method of blinding. Most trials did not select outcome reporting or have incomplete outcome data. In addition, all most of the included trials described the inclusion criteria or diagnose criteria, but one of these(El-Bayoumi[19] 2011) do not mention exclusion criteria.

### 3.3 Network meta-analysis results NMA results

We depicted a network graph of 15 kinds of therapies for GBS. The graph were made by R 3.6.1 software and visNetwork package.

#### Forest graph

There were 22 studies using Disability grade change after 4 weeks as outcome measure, including 13 treatment options, meanwhile, there were 23 studies using rates of improvement by≥1 grades after 4 weeks as outcome measure, including 10 treatment options. The forest figures on results of network pooled comparisons of the clinical effectiveness rate of each therapies and their calculated ranking probabilities were shown in Fig 4 and Fig 5, the forest figure references were Placebo, PE and IVIg.

**Figure 2.**
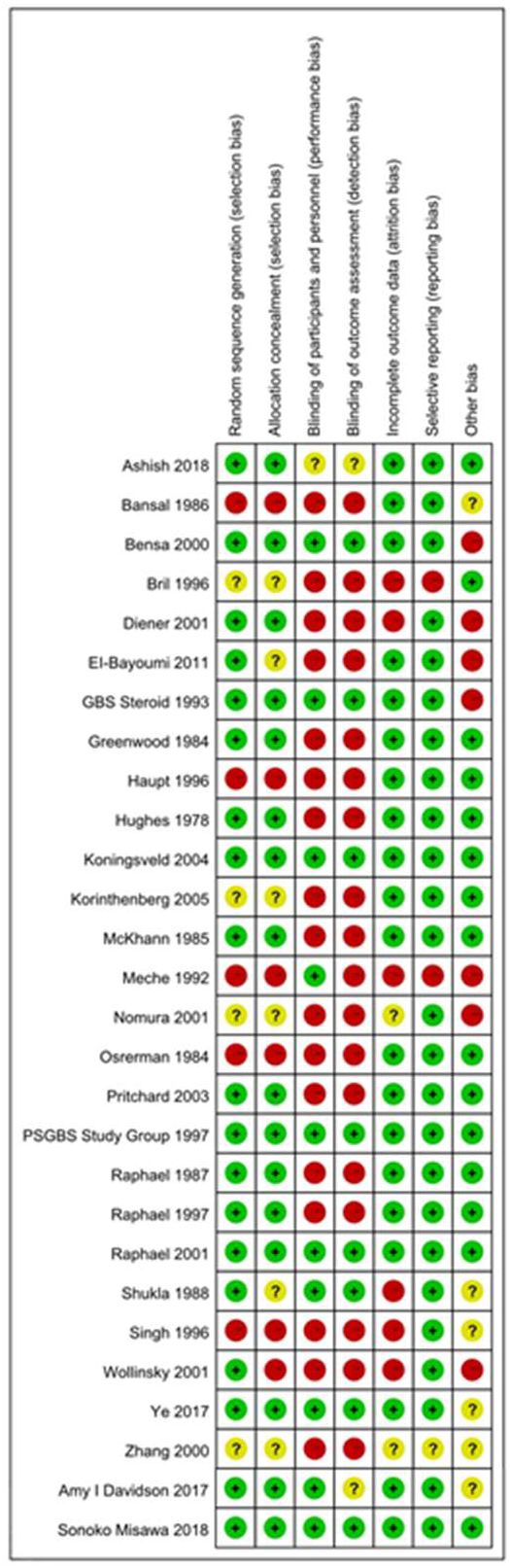
Risk of bias graph.

**Figure 3.**
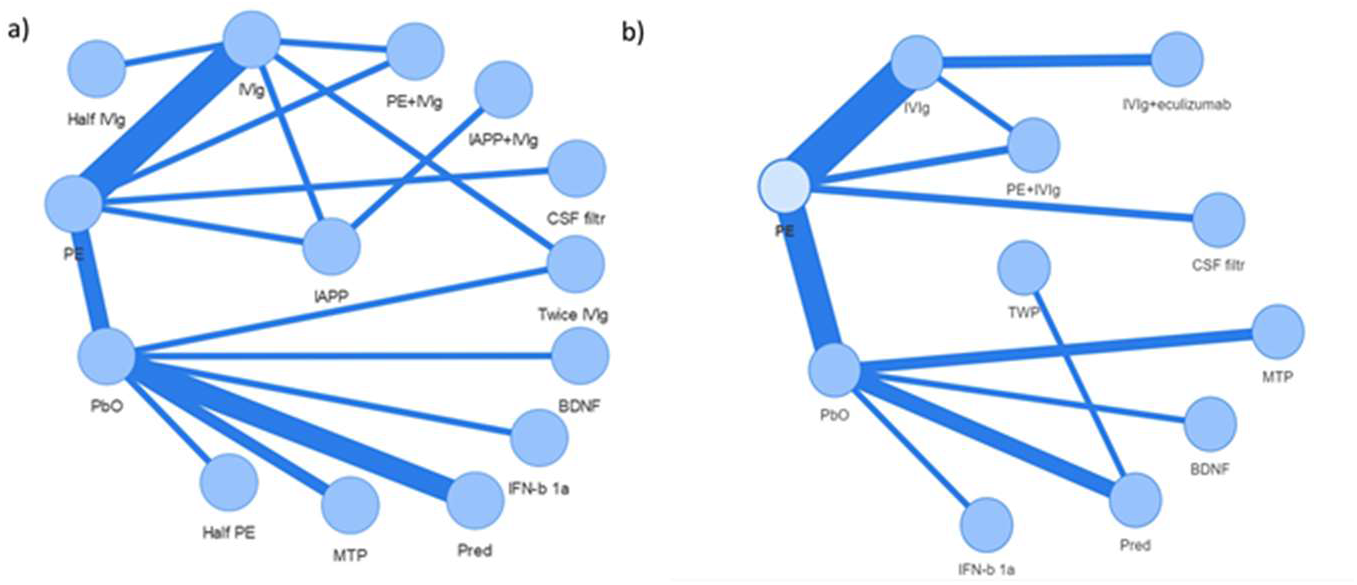
Network graph for therapies of GBS. Note: Line thicknesses correspond to the number of trials used for comparisons. MTP: Methylprednisolone, PbO: Placebo, Pred: Prednisolone, BDNF: Brain-derived neurotrophic factor, CSF filter: Cerebrospinal fluid filtration, PE: Plasma exchange, IVIg: Intravenous immunogloblin 0.4–0.5 g/kg daily for 4–6 days, PE+IVIg: PE followed by IVIg, IAPP+IVIg: Immunoabsorption followed by IVIg, IAPP: Immunoabsorption plasmapheresis, Twice IVIg: 1 g/kg for 2 days, Half IVIg: 0.4 g/kg for 3 days, Half PE: Half course of PE, TWP: Tripterygium Wilfordii Polyglycoside, IVIg+eculizumab: Intravenous immunogloblin 0.4 g/kg daily for 5 days+ eculizumab 900mg once a week for 4 weeks a):trials with outcome ① of Disability grade change after 4 weeks. b): trials with outcome ② of the rates of improvement by ≥1 grades after 4 weeks.

**Figure 4.**
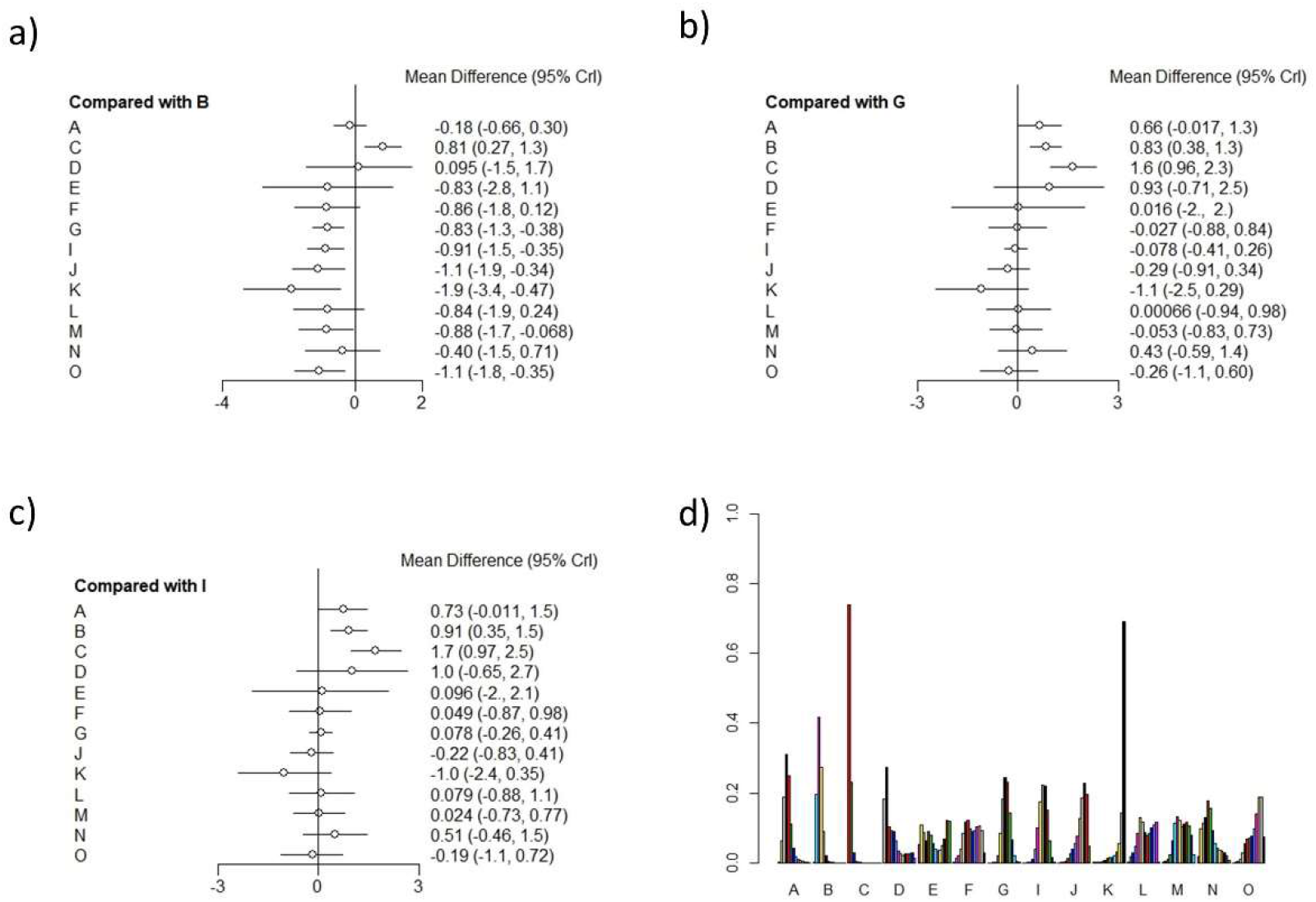
Forest graph and ranking probability graph on outcome ①: Disability grade change after 4 weeks. A: methylprednisolone, B: placebo, C: prednisolone, D: IFNb –1a, E: brain –derived neurotrophic factor, F: CSF filtration, G: PE, I: IVIg 0.4–0.5 g/kg daily for 4–5 days, J: PE followed by IVIg, K: Immunoabsorption followed by IVIg, L: Immunoabsorption, M: IVIg 1 g/kg daily, N: IVIg 0.4 g/kg/day for 3 days O: Half-times of PE a), b) and c): Relative effect comparing with placebo group, PE, and IVIg. d): Ranking probability graph.

**Figure 5.**
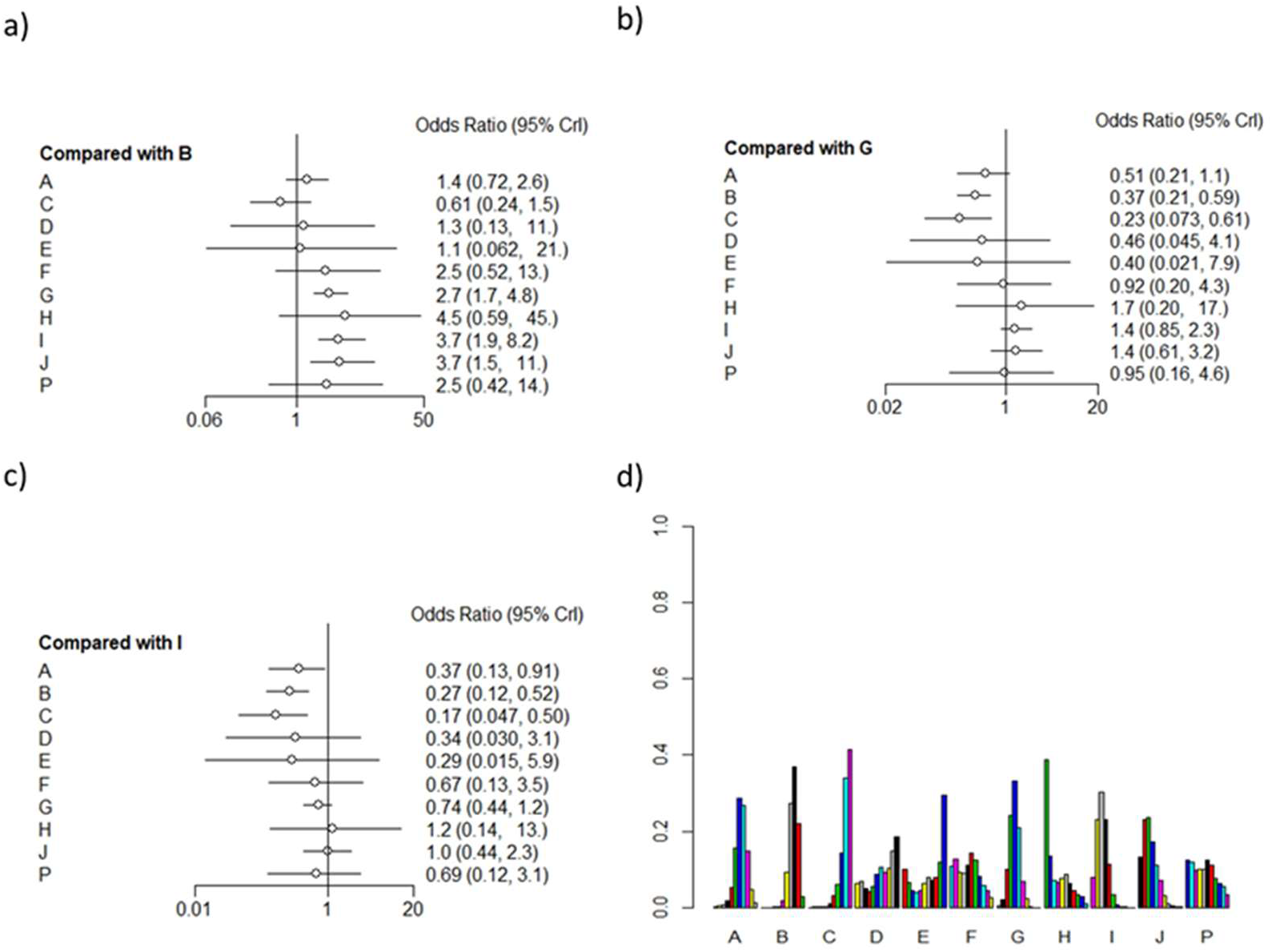
Forest graph and ranking probability graph on outcome ②: rates of improvement by *≥*1 grades after 4 weeks. A-G, I-J: Same with Fig 3. H: Tripterygium Wilfordii Polyglycoside, P: IVIg+eculizumab a), b) and c): Relative effect comparing with placebo group, PE, and IVIg. d): Ranking probability graph.

In the assessment on efficiency of treatments for GBS, PE, IVIg, PE followed by IVIg, immunoabsorption followed by IVIg, IVIg 1 g/kg daily for 2 days, Half-course of PE) were observed to be significantly effective in treating GBS. (outcome①: PE MD = –0.83, 95%Crl[−1.3,−0.38]; IVIg MD = –0.91,95%Crl[−1.5,−0.35]; PE followed by IVIg MD = –1.1, 95%Crl[−1.9,−0.34], immunoabsorption followed by IVIg MD = –1.9, 95%Crl[-3.4,−0.47], IVIg 1 g/kg daily MD = –0.88, 95%Crl[−1.7,−0.068], 2 times of PEs MD = –1.1, 95%Crl[−1.8,−0.35] outcome ②:PE OR = 2.7,95%Crl[1.7,4.8], IVIg OR = 3.7,95%Crl[1.9,8.2], PE followed by IVIg OR = 3.7,95%Crl[1.5,11.0]). Both PE and IVIg were available for GBS (outcome ①: PE MD = –0.83, 95%Crl[−1.3,−0.38]; IVIg MD = –0.91,95%Crl[−1.5,−0.35];outcome②: PE OR = 2.7,95%Crl[1.7,4.7], IVIg OR = 3.6,95%Crl[1.9,8.0])and all kinds of corticosteroids were be indicated no significant efficiency for GBS (outcome①: MTP MD = –0.18, 95%Crl[−0.66,0.30]; Pred MD = 0.81,95%Crl[0.27,1.3];outcome ②: MTP OR = 1.4,95%Crl[0.72,2.6], Pred OR = 0.61,95%Crl[0.24,1.5]). We transferred the base treatment of forest graph for PE and normal dose of IVIg(Fig 4-b),c) Fig 5-b),c)), we could find there were no other therapies being more effective with significant difference. We compared different doses of PE and IVIg (IVIg 0.4–0.5 g/kg daily for 4–5 days, 4–5 times of PE, IVIg 1 g/kg daily, IVIg 0.4 g/kg/day for 3 days, 2 times of PE) and found no significant difference between them. For other kinds of therapies, such as IFNb - 1a, brain - derived neurotrophic factor, CSF filtration, Tripterygium Wilfordii Polyglycoside, IVIg +eculizumab (900 mg once a week) and IVIg 0.4 g/kg/day for 3 days, had no significant difference with placebo.(outcome①: IFNb - 1a MD = 0.095, 95%Crl[−1.5,1.7]; BDNF MD = –0.83,95%Crl[-2.8,1.1]; CSF filtr MD = –0.86, 95%Crl[−1.8,0.12], IVIg 0.4 g/kg/day for 3 days MD = –0.4, 95%Crl[−1.5,0.71] outcome ②: IFNb - 1a OR = 1.1,95%Crl[0.13,11.0], BDNF OR = 1.1,95%Crl[0.056,19.0], CSF filtr OR = 2.5,95%Crl[0.49,12.0], TWP OR = 4.6,95%Crl[0.6,47.0], IVIg+eculizumab OR = 2.5, 95%Crl[0.42,14.0]). Regarding to the improvement for GBS among PE, IVIg, and Corticosteroid, the three most conventional treatments, IVIg was the most helpful one (compared with PE (MD 0.073[−0.26,0.41],with methylprednisolone 0.72[−0.01,1.5],with prednisolone 1.7[0.96,2.5])), but there was no significant difference between PE and IVIg. The efficacy of the two hormones was lower than that of PE and IVIg. (outcome①:MTP VS PE 0.66:[−0.017,1.30] Pred VS PE 1.6 [0.96,2.3] MTP VS IVIg 0.73[−0.011,1.5] Pred VS IVIg 1.7[0.97,2.5] PE VS IVIg 0.078[−0.26,0.41] outcome②:MTP VS PE 0.51:[0.22,1.1] Pred VS PE 0.22[0.075,0.62] MTP VS IVIg 0.38[0.13,0.92] Pred VS IVIg 0.17[0.048,0.50] PE VS IVIg 0.74[0.44,1.2]).

#### Ranking probability

A clustered ranking plot was generated and presented NMA results visually. To better understand the results, the ranking graph was calculated to evaluate the ranking probabilities of all medications on the outcomes. Results were presented in Fig. 4 d) and Fig. 5 d). As suggested by ranking probabilities of outcome ①, immunoabsorption+IVIg had biggest possibility to be a best treatment(P = 0.6), and Half-times of PE, PE followed by IVIg were also likely to be the best treatment. For outcome②, TWP had biggest possibility to be a best treatment(p = 0.4), and IVIg 0.4–0.5 g/kg daily for 4–5 days, PE followed by IVIg followed. For most probability to be worst treatment, in out come① was prednisone, IFNb - 1a came second, in out come② was prednisone, IFNb - 1a and BDNF followed.

#### League Table Heatmap

We use nma.league() Function in BUGSnet package to produce League Table Heatmap. The map would show comparison results of each therapy clearly. (Fig. 6 and Fig. 7)

**Figure 6.**
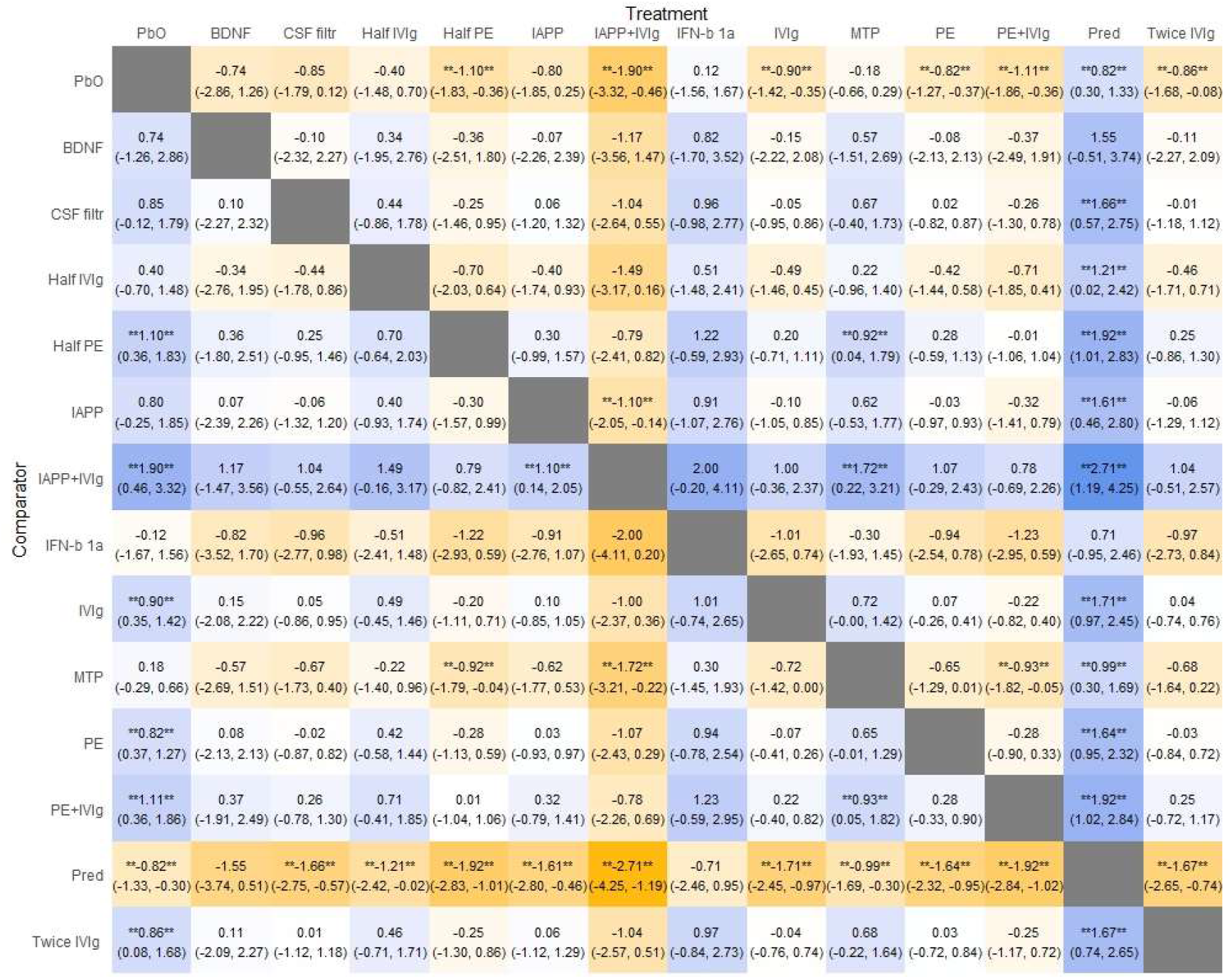
League Table Heatmap Result for outcome ① Legend: The values in each cell represent the relative treatment effect (and 95% credible intervals) of the treatment on the top, compared to the treatment on the left. A double asterisk indicates statistical significance

**Figure 7.**
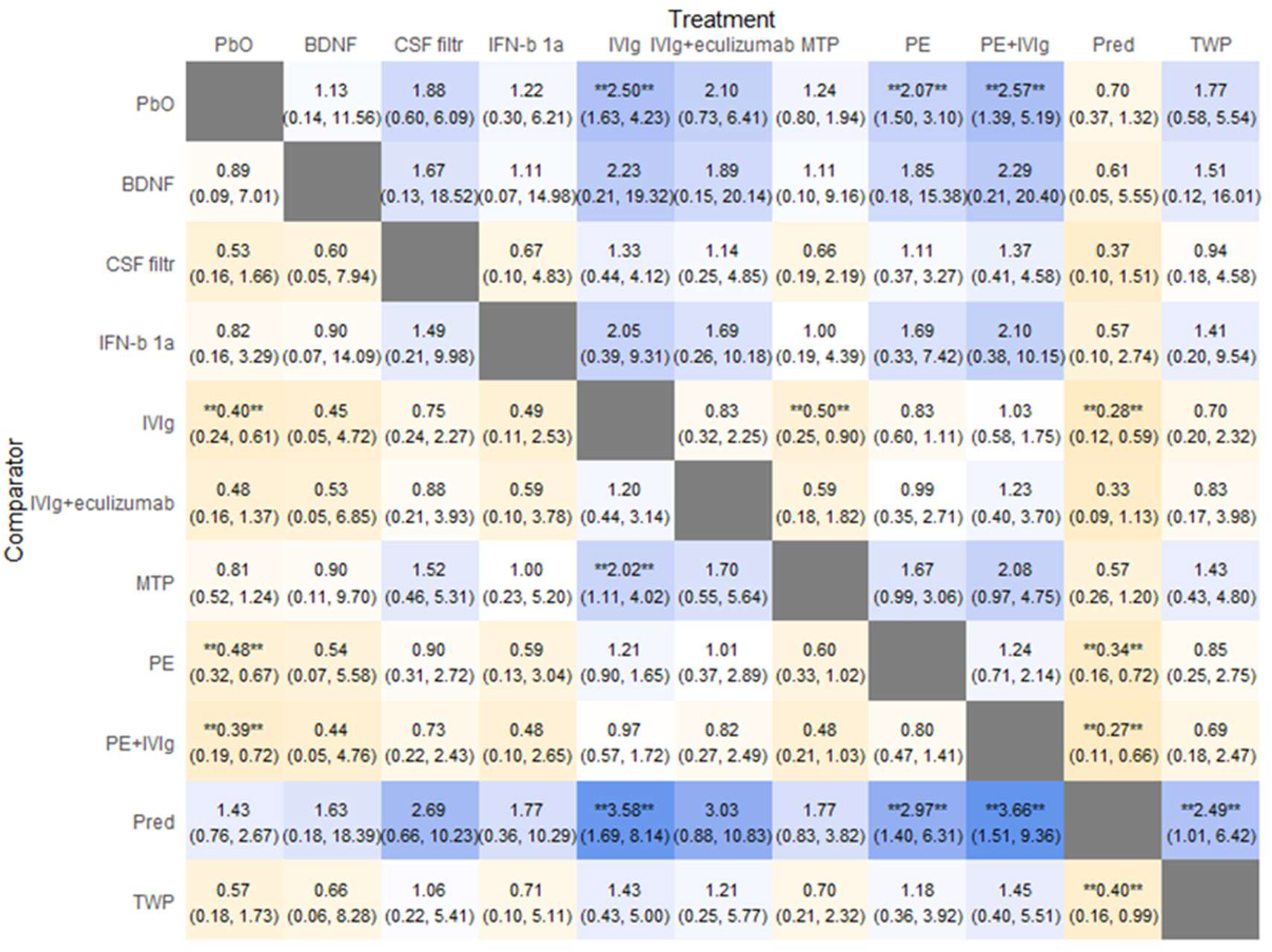
League Table Heatmap Result for outcome ②

#### Gelman-Rubin-Brooks plot

As we carried network meta-analysis based on a bayesian hierarchical framework, we should confirm our simulations have resulted in the convergence of the algorithm, which represented the stability of our results. The plot showed well convergence of the algorithm(Fig 8.).

**Figure 8.**
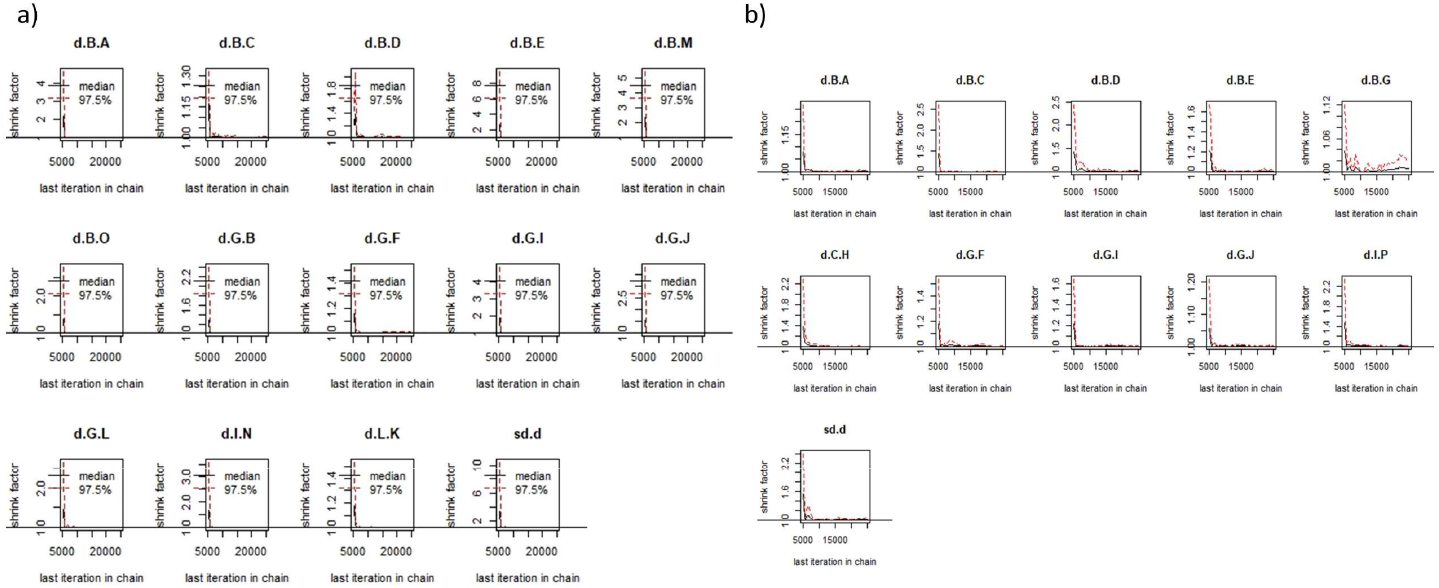
Gelman-Rubin-Brooks plot for NMA. a)Plot for trials with outcome ① of Disability grade change after 4 weeks. b): Plot for trials with outcome ② of the rates of improvement by ≥1 grades after 4 weeks.

#### The order of SUCRA value

If a treatment always ranks first, then SUCRA = 1, and if it always ranks last, it will have SUCRA = 0[20]. We use the SUCRA function in dmetar package to calculate SUCRA Score and ordered it in descending order.(Fig 9)We found Conventional dose of IVIg, PE followed by IVIg were in the front of therapies queue. The SUCRA score of PE was lower by IVIg in graphs. Corticosteroids on two outcomes were in the bottom. For the outcome①, BDNF, CSF filtration, PE, Conventional dose of IVIg, Immunoabsorption, IVIg 1 g/kg daily got a similar SUCRA value, which could from the side indicated that there may be no significant difference in efficacy between these treatments.(Fig. 9 a)). For the outcome②, PE got a similar SUCRA value with IVIg+eculizumab.

**Figure 9.**
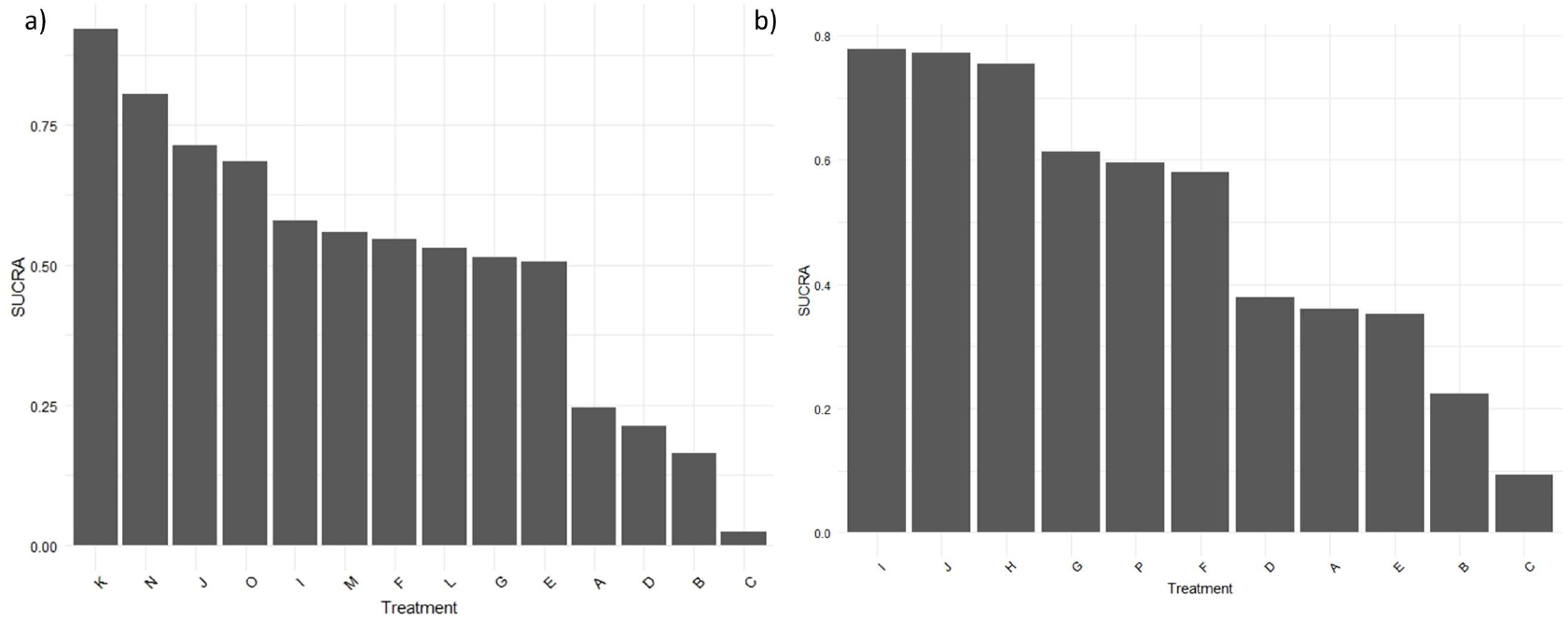
SUCRA value of the treatments(descending order) a):SUCRA value in outcome①:A-G, IJ: Same with Fig 3. b): SUCRA value in outcome②: A-J, P: Same with Fig 4.

### 3.4 Consistency analysis and heterogeneity test

We used I^2^ for consistency checks direct results, From the Fig 10. and Fig 12, almost all of the I^2^ were under 50% which means the heterogeneity of the direct NMA was in a lower rage, the results of direct evidence of the NMA were reliable. We used the fixed effect model for meta-analysis.

**Figure 10.**
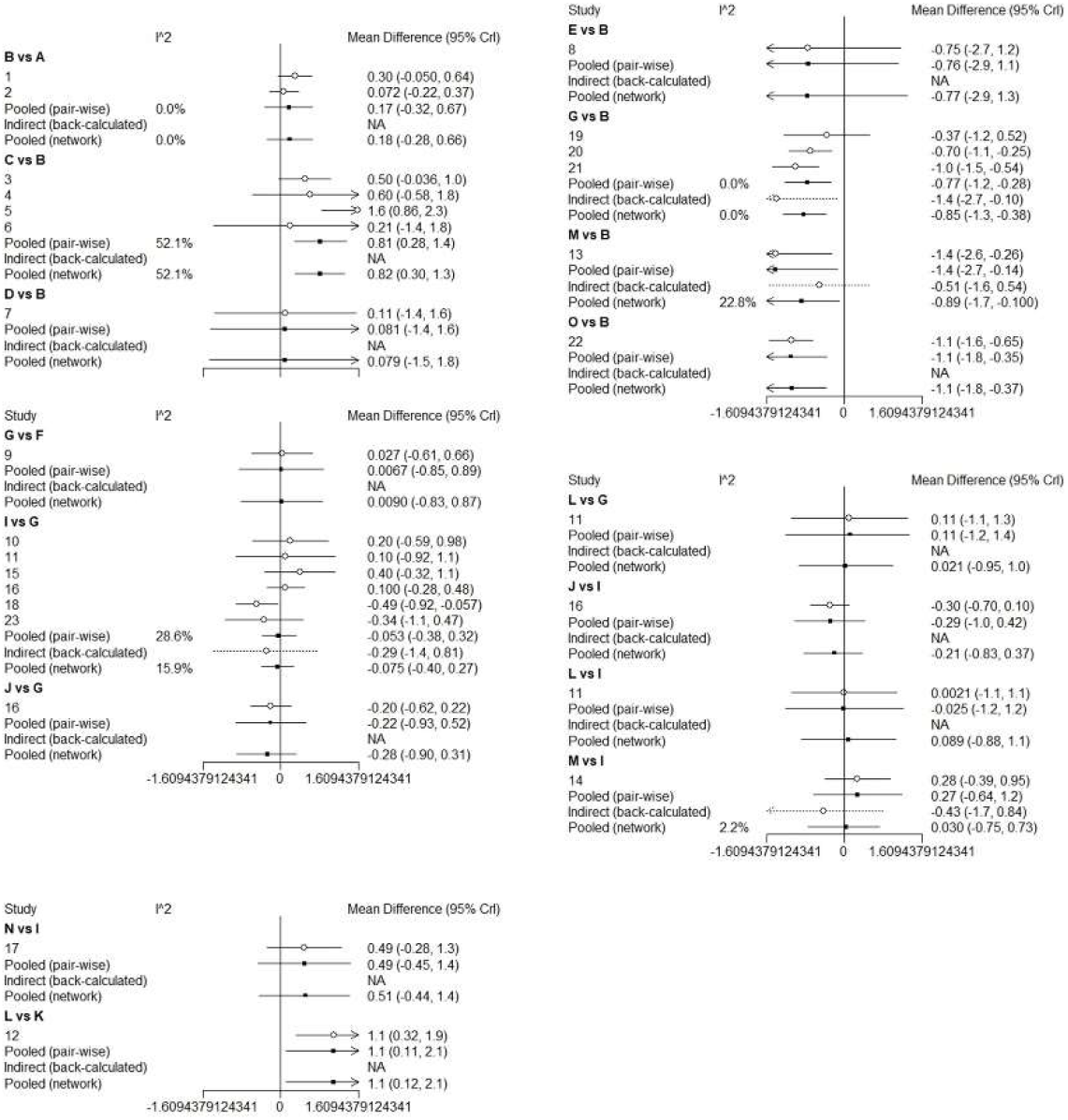
Consistency analysis of outcome ① A-G, I-J: Same with Fig 3.

**Figure 11.**
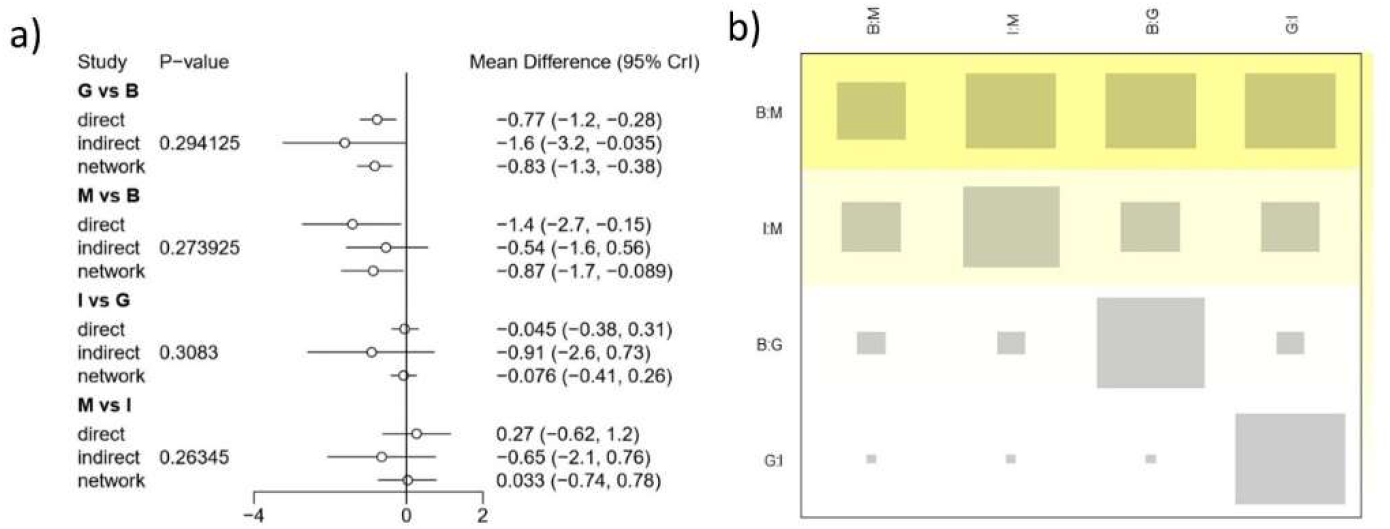
heterogeneity test of outcome ① A-G, I-J: Same with Fig 3.

**Figure 12.**
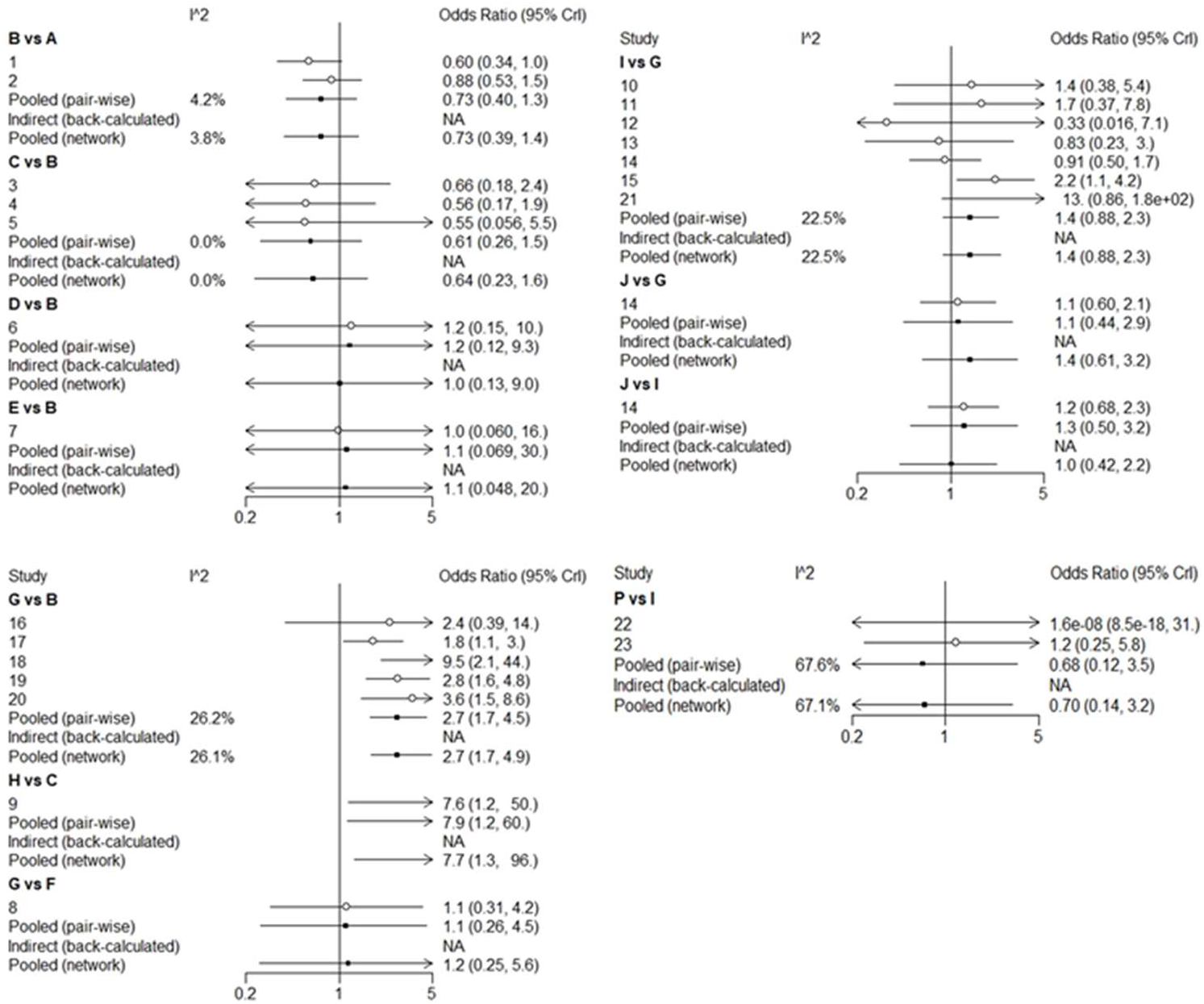
Consistency analysis of outcome ② A-J: Same with Fig 4.

The node-splitting method and its Bayesian P value was used to report the inconsistency of our results between direct and indirect results. For the majority of our results, the confidence intervals from direct and indirect evidences were in consistent, with minor differences. In the inconsistency checks, we found that there were some heterogeneity between the four groups in outcome①, (G VS B,M VS B,I VS G,M VS I), as there was no obvious heterogeneity in outcome②. To further determine the heterogeneity, we used netheat diagrams in the netmeta package for heterogeneity analysis. The results showed that the heterogeneity was within the acceptable range. We further used the direct.evidence.plot function in the dmetar package (from github) to analyze the sources of direct and indirect evidence, and the results showed that the three groups of results (G VS B,I VS G,M VS I) were dominated by direct evidence. The results of M VS B were mostly based on indirect evidence, but the results of network evidence were consistent with its direct evidence, as we thought direct evidence had more credibility. Based on the above results, we believed In outcome①, there was no significant heterogeneity in the network meta analysis which may influence the result significantly. In the outcome②, we did not find groups with significant heterogeneity. Finally, we could see from the direct.evidence.plot that most of the comparison results in this study are obtained through indirect comparison. Since the indirect results of the meta were calculated based on the bayesian algorithm, they still needed to be verified by a large number of direct comparisons.

### 3.5 Publication bias

Funnel plots were used to measure the publication bias. The funnel plot of the improvement in Disability grade change after 4 weeks, and the rates of improvement by≥1 grades after 4 weeks showed potential publication bias of the included RCTs (Fig. 14). It can be seen from the funnel plot that almost all the studies fall within the funnel and the two sides of the funnel were basically symmetrical, so the possibility of publication bias in this study was small.

**Figure 13.**
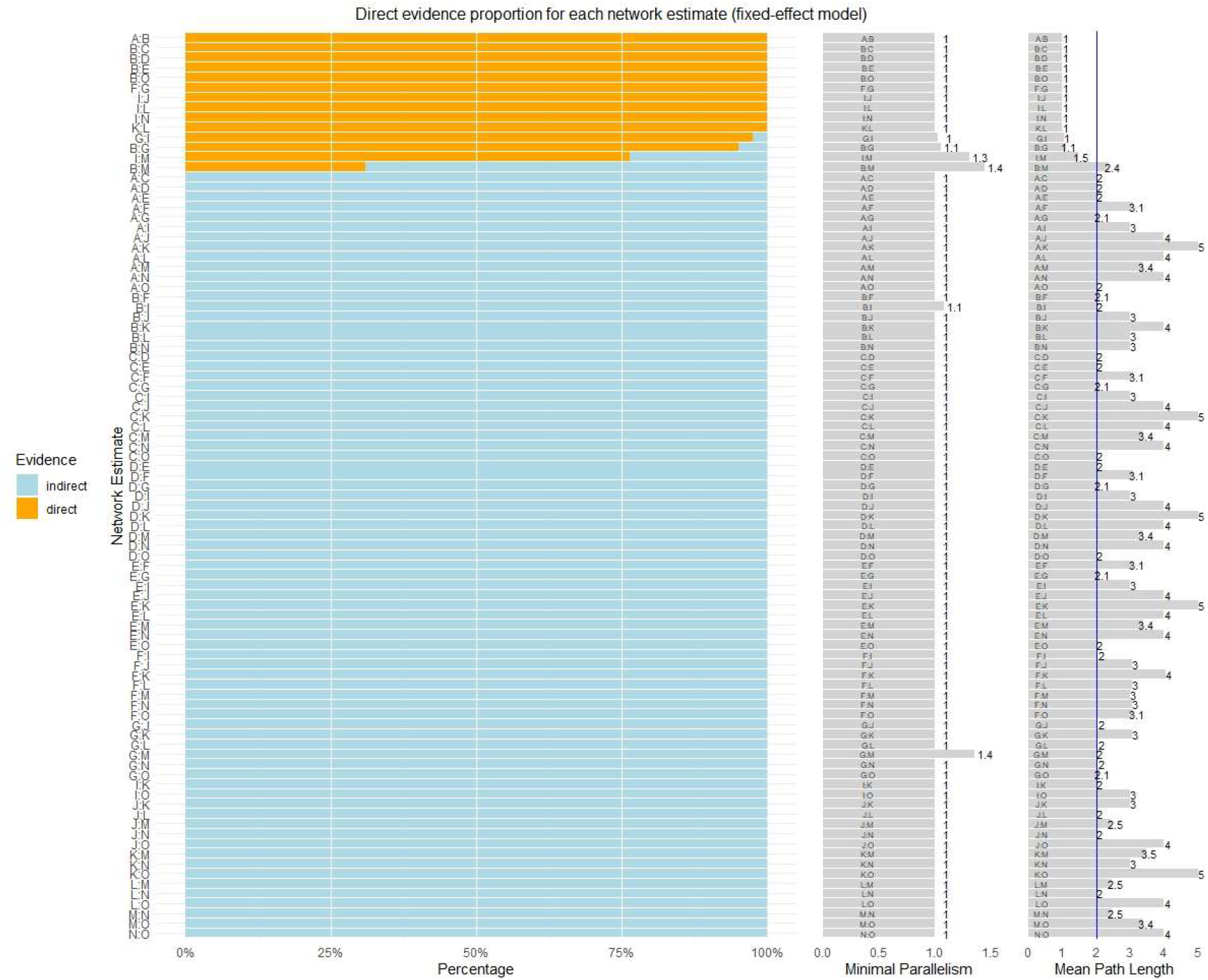
Direct evidence plot for outcome ① A-G, I-J: Same with Fig 3.

**Figure 14.**
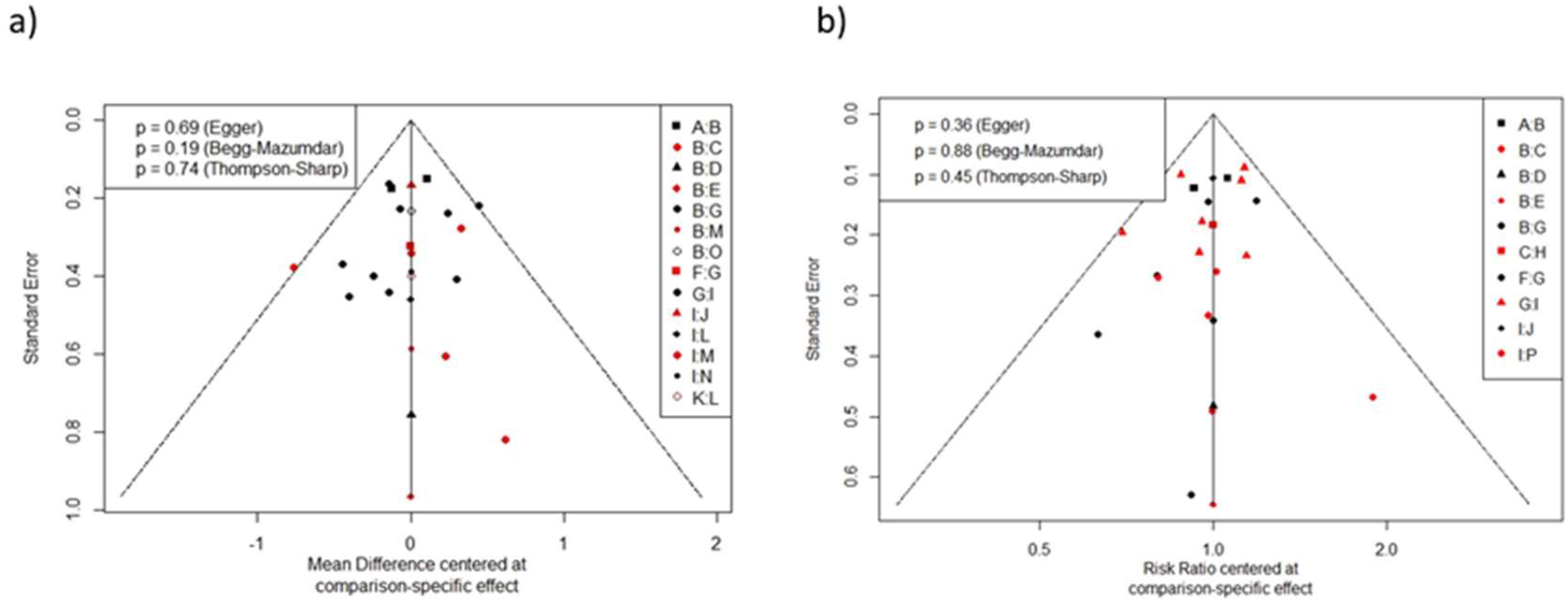
Funnel plots for outcome ① and outcome ②

## 4. Discussion

### On NMA results

PE and IVIg have been widely used in clinical practice to alleviate GBS[10, 12], and recommended by the European guidelines[21, 22] rather there were some evidence indicating corticosteroids had no significant efficiency for GBS [9]. In our studies, PE and IVIg as therapies for GBS are not weaken to every other kinds of therapies. Considering that two therapies are conventional treatments for GBS, our NMA helps to confirm their effectiveness in clinical practice[23–27]. Immunoabsorption+IVIg had highest possibility for 1st rank for the improvement Disability grade change after 4 weeks in a small sample, which may infer the better efficiency on GBS treatment. However, the number of patients included was only 21, the sample size was insufficient, moreover, the comparison with PE and IVIg is indirect, so its efficacy still needs to be further verified with a large sample research. Corticosteroids were not effective as reported before[28, 29]. IVIg+eculizumab was a relatively new option for GBS, but in NMA, it didn’t show advantages comparing with IVIg and PE. The samplesize of IVIg+eculizumab was also insufficient. For Outcome①, BDNF, CSF filtration, PE, Conventional dose of IVIg, Immunoabsorption, IVIg 1 g/kg daily got a similar SUCRA value, so, maybe some unconventional therapy should be given more attention on, A larger sample study may yield positive results.

In the aspects of outcome② for the GBS, we can find that the results of PE and IVIg were similar to that of outcome①, both of which had good therapeutic effects, but there was no significant difference between the two. The rank and SUCRA of IVIg are better than those of PE. Tripterygium Wilfordii Polyglycoside showed its advantage in treatment results. However, with insufficient sample size and unclear adverse reactions, further researches on TWP were needed.

The results of Outcome① and outcome② are basically the same, which enhanced the reliability of the study. For another hand, However, many of these therapies had only one of the included studies reported the efficacy, and most of the comparison results were obtained through indirect comparison. Therefore, whether the conclusion is accurate or not, more direct comparison and indepth study are still needed.

In terms of different doses and treatment courses of PE and IVIg, the rank obtained by half course of PE is better than that of PE and IVIg. However, we could see from the network that the comparison object of half course of PE was placebo, the indirect result with small samples can not be conceived. At the same time, for IVIg 1g/kg over 2 days, we could see that the patients of two trials comparing with IVIg 1g/kg over 2 days were different. One was for children, the other was for adults. And the control groups were IVIg and placebo respectively. Although we calculated that the indirect comparison of IVIg 1g/kg over 2 days was superior to the IVIg, we thought that direct result—conventional IVIg was better than IVIg 1g/kg over 2 days more conceivably. In general, there were no significant difference between all of different doses and times of PE and IVIg.

For the immumoabsorption, in two essays[30, 31], we could find it shown equally favorable results in acute GBS comparing with PE or IVIg. On the clinical adverse reaction, treatment-related adverse reactions were, however, fewer in the immunoglobulin group compared to the other groups[31]. Immumoabsorption also has been used successfully in myasthenia gravis[32, 33], which is also an autoimmunity disease. So, we thought it could be an complement method for GBS besides PE and IVIg.

Combination therapy has been somewhat controversial for guillain-barre[21, 22]. PSGBS Group 1997 carried out a result of no significant difference of combined treatment of PE and IVIg comparing with PE or IVIg. In our studies, we put its data into the network meta, we would found the PE+IVIg got a higher rank than IVIg, even if the advantage is slightly. As for Immunoabsorption followed by IVIg, we could find it got highest rank and SUCRA value in outcome①. Haupt [30] showed that immumoabsorption followed by IVIg may lead to a more rapid recovery of patients suffering from GBS when compared to those patients treated by immumoabsorption or PE alone. Consequently, we thought the effect of combination therapy, especially for PE followed by IVIg, Immunoabsorption followed by IVIg needs to be further researched.

### On Consistency test and heterogeneity test

In the meta-analysis, we adopted the fixed effect model wholly. Most of I^2^ was less than 50%, while only B(Placebo) VS C(Prednisolone) was 52.1%. However, all the studies showed that B was better than C, so we insisted on adopting the fixed effect model. In terms of the heterogeneity analysis of indirect and direct evidence(Fig 11.), we can see P>0.1 through the node cracking method, while the yellow and red color of netheat plot is not very deep, and there is no inverted blue square, which indicated heterogeneity in a low level. In this study, NMA was mainly carried out through bayesian algorithm and gemtc package, but the gemtc package could not draw the heterogeneity heatmap and funnel figure, so we used netmeta package to draw the heterogeneity netheat plot and funnel figure. Previous literature showed that the results from netmeta package were very similar with the results from a fixed effect Bayesian analysis[34], so the plot and funnel figure could be transplanted from using netmeta package as a reference for heterogeneity analysis and publication bias. We believed that the heterogeneity is under control, so the conclusion obtained through indirect comparison in this paper has certain reference value. In additional, we use BUGSnet package to get League Table Heatmap to show all of NMA analysis results. BUGSnet requires that the user have installed Just Another Gibbs Sampler (JAGS) on their computer, it’s result in accordance with getmc package highly[16]. At the same time, for the indirect evidence conclusion of well efficiency treatments, such as IVIg followed by PE, tripterygium wilfordii polyglycosides, etc.., we should also carry more clinical trials for affirming the efficacy of them.

Previous clinical studies confirmed the efficacy of PE and IVIg, and meta-analysis further confirmed and elucidated the efficacy. We demonstrated the effectiveness of these two approaches from a larger range of treatment options. In the treatment of corticosteroid, the results demonstrated that in a larger context, the efficacy of corticosteroid in treating guillain-barre syndrome was limited. Compared with RCT, our results indicated a more integral conclusion, and the accumulation of evidence from randomized and cohort clinical trials could lead to a more convincible conclusion on the efficacy of therapies on GBS.

### Limitation

The primary limitation of this analysis was the limited sample size of involved therapies and subjects, especially for some treatments with well results, such as combination treatments of IVIg+PE and IVIg+immumoabsorption. In addition, there were some different age groups in the included researches. In Korinthenberg[35] 2005, the main patients of GBS were children, others were adults. At the same time, since most of the treatment methods of GBS are non-drug treatments, the implement of blind methods will be limited to some extent, so the number of blind methods(36% using blind) in the included literatures is relatively small. Besides, the inclusion criteria and disability scale were not identical. In addition, it has been suggested that a more unified GBS diagnostic criteria and more unified evaluation of curative effect should be established to assess GBS accurately. Larger and well-designed clinical trials on the efficacy of GBS on patients should be needed for further investigation.

### Conclusion

In conclusion, we observed in our meta-analysis that PE or IVIg had a significant efficiency for GBS patients. They can help to improve activities of daily living. Different doses of IVIg or PE, combination of PE and IVIg had no significant difference with PE and IVIg alone in Conventional dose. The effect of IVIg+PE, IVIg+immumoabsorption, IVIg+eculizumab, Tripterygium Wilfordii Polyglycoside should be further explored. Corticosteroids have no significant effect on GBS. Larger clinical trials on the efficacy of therapies with GBS are needed for further investigation.

**Table 1:**
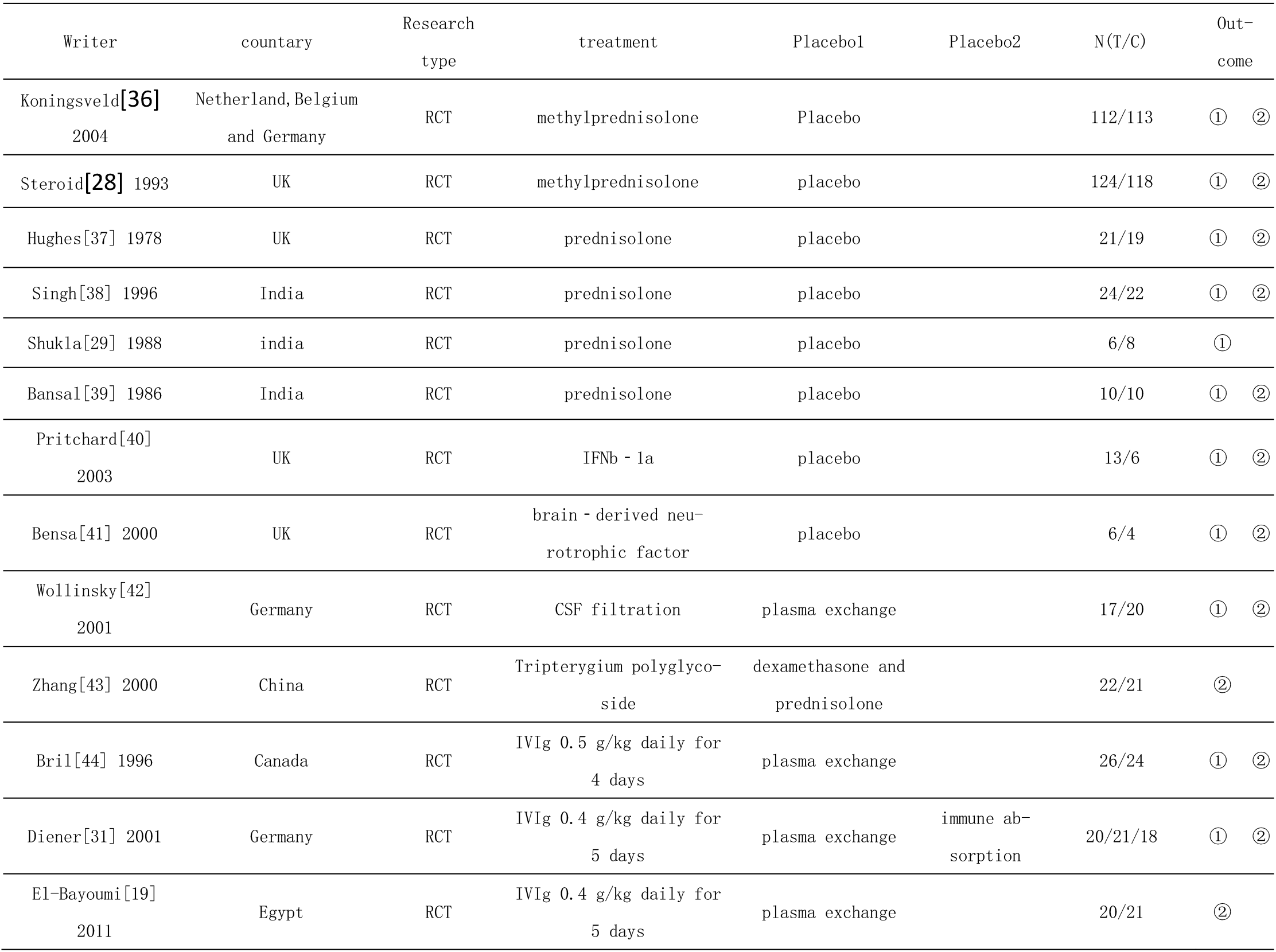
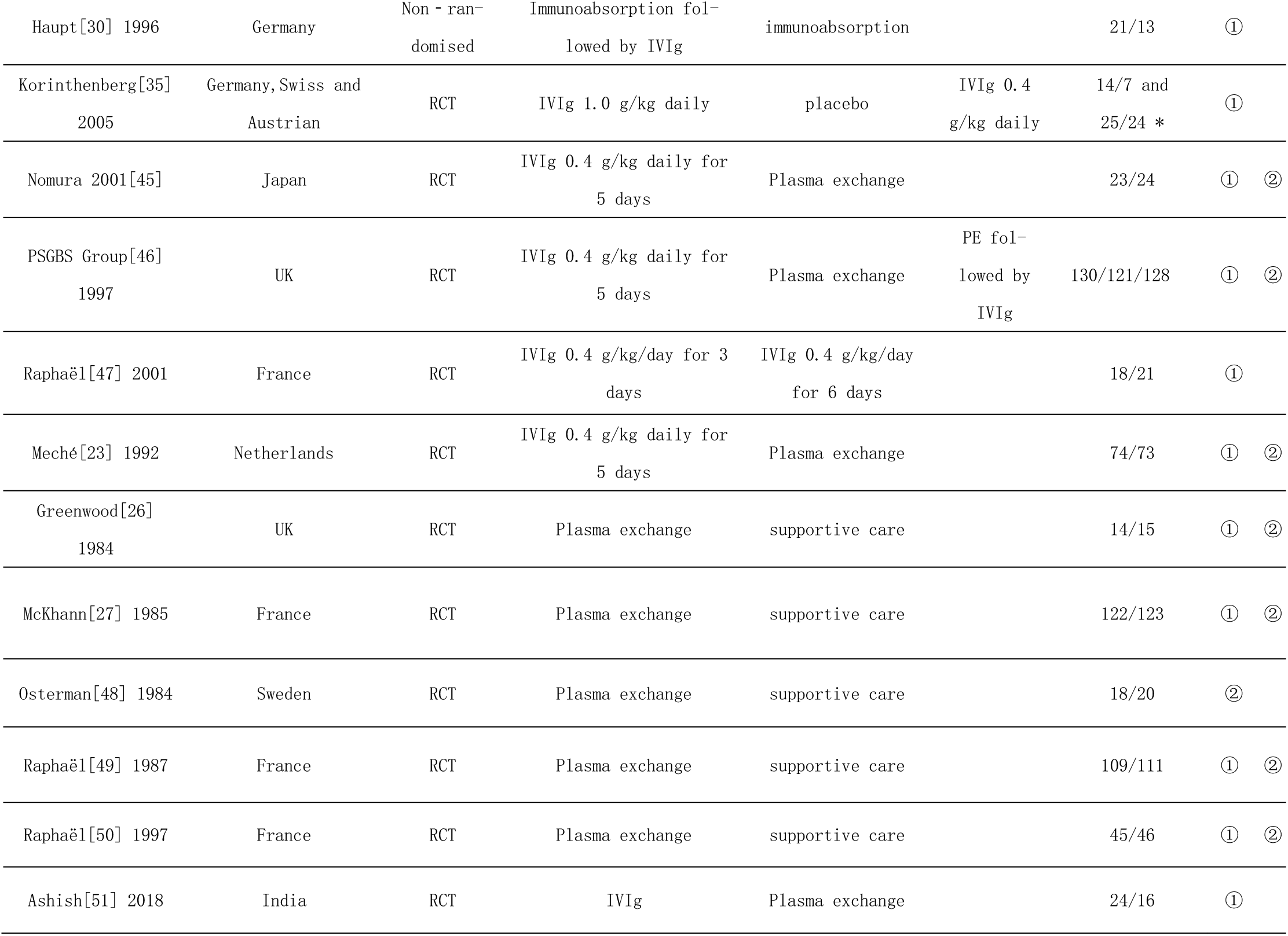
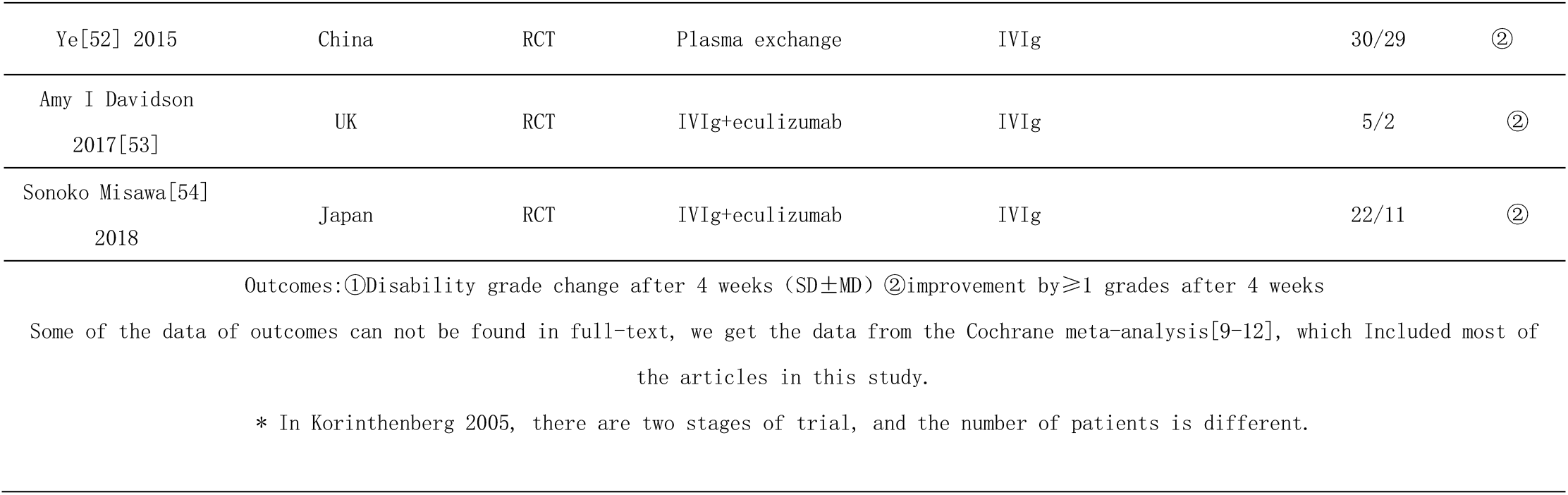
Characteristics of enrolled trials

## Data Availability

All codes and data are available. The data URLs will be open after accepted.

https://github.com/jingfenglin/data-for-guallian-barre

## Abbreviations

NMA: network meta analysis
GBS: Guillain-Barre Syndrome
MTP: Methylprednisolone
PbO: Placebo
Pred: Prednisolone
BDNF: Brain-derived neurotrophic factor
CSF filter: Cerebrospinal fluid filtration
PE: Plasma exchange
IVIg: Intravenous immunogloblin 0.4–0.5 g/kg daily for 4–6 days
PE+IVIg: PE followed by IVIg
IAPP+IVIg: Immunoabsorption followed by IVIg
IAPP: Immunoabsorption plasmapheresis
Twice IVIg: 1 g/kg for 2 days
Half IVIg: 0.4 g/kg for 3 days
Half PE: Half course of PE
TWP: Tripterygium Wilfordii Polyglycoside

## Declarations

### Ethics approval and consent to participate

Not applicable

### Authors’ contributions

J.Lin and Q.Gao had full accessed to all the data in the study and takes responsibility for the integrity of the data and the accuracy of the meta analysis. J.Lin, K.Xiao and Q.Gao designed the study and determined the content. J.Lin determined the retrieval scheme, collected the data, performed the statistical analysis and drafted the manuscript. Q.Gao supervised the study, doublechecked the statistical analysis. D.Tian and Y.Hu screened articles. Z.Han revised the manuscript. All authors have read and approved the manuscript.

## Acknowledgements

Thanks to Monkey Brother in Zhihu, who shared his NMA experience. Thanks to Richard AC Hughes and his team’s meta-analysis results published in Cochrane Library.

## Appendix

Appendices

